# Perspectives of young people, family carers and voluntary sector staff on help-seeking for mental health difficulties in a rural region of the United Kingdom - a qualitative study

**DOI:** 10.1101/2024.01.04.24300825

**Authors:** Sheri Oduola, Emma Coombes, Jo Hodgekins, Andy Jones

## Abstract

**Purpose:** To explore key stakeholders from rural communities’ perspectives of help-seeking for mental health difficulties.

**Methods/design:** Semi-structured interviews and a focus group were conducted with participants living in the East of England, UK. Thematic analysis was used to organise and analyse the data.

**Results:** Nine participants were recruited. Participants described barriers and facilitators of help-seeking. Barriers included: lengthy waiting times, socioeconomic means, poor therapeutic relationships, insufficient treatment duration, poor transition between services, high staff turnover, and limited investment. Facilitators included: family involvement/support, school nurses, shorter delays in accessing specialist services such as eating disorder services, and the offer of family therapy. Suggestions for improving future help-seeking and pathways to care, included: peer-support for young people, enhanced support for carers, early intervention and raising awareness of mental health.

**Originality:** Our qualitative approach meant we were able to explore in detail the existence and mechanisms underlying varied barriers and facilitators of care pathways amongst our sample. This allowed us to report rich findings and recommendations for clinical practice and future research work.

**Practical implications:** Training young people to support other young people, and identifying mental health champions in rural communities could reduce distress and ameliorate pressures on the health service.

## INTRODUCTION

Mental health disorders such as depression, anxiety, and psychoses are relatively common and debilitating (WHO, 2018). The risk of these disorders tends to increase during and after puberty (Lewinsohn et al., 1998), and they are burdensome to young people because the severe impairment can extend beyond the experience of the distress and symptoms. The burden of these mental health disorders includes unemployment, educational difficulties, discrimination, and stigma (Ford et al., 2021; WHO, 2018). Timely access to specialist mental healthcare has been shown to reduce the likelihood of poor clinical outcomes (Bhui et al., 2014) and bring relief to families and society (Onwumere et al., 2018).

Whilst the incidence of mental health disorders is comparable between people living in rural and urban areas in the UK (Kirkbride et al., 2016), pathways to care (PtC) for mental health problems among young people are well studied yet often in urban populations (Allan et al., 2020; Hodgekins et al., 2017; MacDonald et al., 2021). For those living in remote areas, access to specialist treatment is limited (Morales et al., 2020). There are several reasons for this disparity; research has shown that young people living in rural and remote areas face isolation and difficulties in getting help for their mental health difficulties because of poor transport, a lack of safe spaces to meet, and poor digital connectivity (Allwood, 2021; Costas, 2017). From a service provision perspective, most specialist mental health services are based in city centres, hence the practical challenges inevitably have implications for treatment delays and recovery for rural populations. Further, the impact of shame, stigma, and discrimination in seeking help for a mental health problem (Morales et al., 2020), as well as geographic isolation, reduced access to services and lower socioeconomic status (Andrilla et al., 2018) are significant barriers for rural populations when seeking mental health support. Despite these concerns, there are many gaps in the literature, limiting the development of interventions to overcome the barriers identified. As well as being socially unjust, these inequalities in access to care and treatment are concerning as they can lead to greater life-long disability in future generations of adults.

Young people’s mental health is a priority area in the UK NHS Long-term Plan, which pledges to develop new approaches to supporting young adults aged 16-25 by bringing together partners in health, social care, education and the voluntary sector, through the Integrated Care System (NHS England, 2019). Voluntary organisations play a significant role in care provision that is recognised in the NHS Mental Health Implementation Plan, acknowledging the need for new models of integrated, multi-agency care for young people with mental health difficulties (NHS England, 2019; NHS Providers, 2019). Yet, there is limited research investigating partnership working between the third sector and statutory mental health services. Meanwhile, the real-world implementation of the integrated care system has been slow, hindered by funding crises, insufficient empirical evidence, and poor public engagement. The prolonged period of austerity in the UK has also meant that mental health services remain underfunded (The King’s Fund, 2018). Understanding these issues is important to inform strategies for providing more equitable access to mental healthcare.

In this study, we engaged key stakeholders from rural areas (i.e., young people with lived experience, family carers and voluntary organisation workers) in qualitative interviews and a focus group to explore their help-seeking experiences for mental health difficulties, and the potential interface of voluntary organisations with statutory mental health services. Our research was guided by the question “What are young people’s, family, and voluntary sector’s perspectives on the barriers and facilitators of seeking help for mental health problems in rural areas and how can we improve access to care?”

## MATERIALS AND METHODS

### Study design

A qualitative design was employed to explore young people’s, family carers’, and voluntary organisation worker’s experiences of help-seeking for mental health difficulties. A mix of convenience and purposive sampling were used to identify participants.

### Settings and participants

Participants were recruited from community settings in the East of England region, UK. The region is a mixture of rural and urban areas but consists of sizeable rural populations (Dept. for Environment Food & Rural Affairs, 2021). Recruitment was conducted between November 2021 and February 2022. Our purposeful sampling approach aimed to ensure diversity. Study flyers and promotional materials were sent to community groups, and charitable organisations to advertise the study to their service users and staff. Study flyers were also promoted to young people at the local Sixth form schools/colleges. Those expressing interest were screened based on a set of inclusion/exclusion criteria and those meeting the criteria were consented to take part in the study.

### Inclusion/exclusion criteria

1. Service users – young people (aged 16-25 years) who had experience of seeking help from a statutory (e.g., NHS) mental health service or voluntary organisation for mental health difficulties (e.g., depression, anxiety, or non-organic psychoses), residing in a predominantly rural setting, and who were able to give informed consent to participate in this research.
2. Family carer – People aged over 16 who were informally supporting a young person currently seeking help or having previously sought help from an NHS mental health service or a voluntary organisation, residing in a predominantly rural setting, and who were able to give informed consent to participate in this research.
3. Voluntary organisation staff – People in voluntary organisations (in either paid or voluntary capacities) with experience of supporting young people with mental health difficulties living in predominantly rural areas and who were able to give informed consent to participate in this research.

### Data collection

A semi-structured interview schedule was designed using topic guides, which included exploration of participants’ recognition of the onset of mental health difficulties, experiences of pathway to care, influence of rurality in help-seeking behaviour, the role of the voluntary sector in help-seeking, and potential strategies for improving timely access to care. Interviews were conducted by members of the research team (SO, EC, JH, AJ), who are all experienced researchers with two also being clinicians, (SO, a mental health nurse and JH, a clinical psychologist). Interviews were conducted either face-to-face, or remotely via videoconferencing, and were audio recorded. During the interview, participants were asked to complete a short demographic survey. Two initial interviews (by AJ and SO) were discussed by the research team to check that the interview questions sufficiently identified data relevant to the research questions. The team agreed that the questions were adequate, therefore no changes were made, and both interviews were included in the analysis. For their convenience, three young people participated in one focus group discussion, conducted in their school. The focus group was facilitated by EC and AJ, and the discussions were guided by the above topic guides. All discussions lasted between 30-90mins.

### Data analysis

Thematic analysis was used according to the principles set out by Braun and Clarke (2006)(Braun & Clarke, 2006). Data were transcribed verbatim, initial codes generated, themes searched for, themes reviewed, and themes defined and named. All authors were involved in the data analysis. First, the researchers became familiar with the data by discussing participants responses after each interview during monthly team meetings. Second, one author (EC) transcribed the interviews. Third, generating a coding framework, initial codes and emerging themes were identified by one author (SO) using NVivo 20 (Dhakal, 2022; QSR International Pty Ltd, 2020). A freely available mind-map software, *Wondershare Edraw*, was used to organise the themes as they emerged (Edrawsoft, 2022). To ensure data relating to the codes and themes were not missed, and to maintain rigour, the transcripts and mind-maps were reviewed by all the authors in a team meeting briefing. Finally, all authors were involved in naming the overarching and sub-themes.

### Ethical approval

Ethical approval was obtained from the University of East Anglia, Faculty of Medicine & Health Sciences Research Ethics Committee (Reference: 2021/22-018). All participants were provided with information about the study, offered an opportunity to speak with the researchers prior to taking part, and gave written consent. Participants were given a £25 gift voucher each, to thank them for taking part in the study.

### Patient and Public Involvement

The research team discussed and advertised the study with voluntary organisations and community groups. There was no patient involvement in the design of the study.

## RESULTS

### Sample characteristics

Nine people (4 family carers (FC), 4 young people (YP) and 1 voluntary organisation staff member (VOS)) agreed to take part in the study. Our sample was relatively homogenous; most participants were female, employed or studying, many lived in a village or hamlet, and all were white British. All family carers were aged 46 years and above, and the young people were aged 16-18 years. Table 1 shows the participants’ sociodemographic characteristics. Of note, the family carers and young people in our study were recruited independently, and they did not share familial relationships. No one requested to withdraw from the study, nor did anyone drop out.

**Table 1:** Sample characteristics.

|  | Young people | Family Carers | Voluntary organisation staff |
| --- | --- | --- | --- |
| Individuals interviewed | 4 | 4 | 1 |
| Age band | 4 (16-18 years) | 4 (46+ years) | 26-35 years |
| Gender | 1 Neutral<br>3 Females | 4 Females | Non-binary |
| Ethnicity | 4 White British | 4 White British | White British |
| Employment status | 4 Student | 4 Employed | Employed |
| Area of residence | 1 Hamlet<br>1 Village<br>2 Market town | 1 Hamlet<br>2 Village<br>1 Market town | Village |

### Themes/sub-themes

Our thematic analysis highlighted three overarching themes: (a) pathways to care experiences, which refers to help-seeking attempts when an individual recognises a mental health difficulty, (b) structural/organisational factors, and (c) strategies for improvement, and thirteen sub-themes (Figure 1). Barriers and facilitators for each theme are discussed.

**Figure 1:**
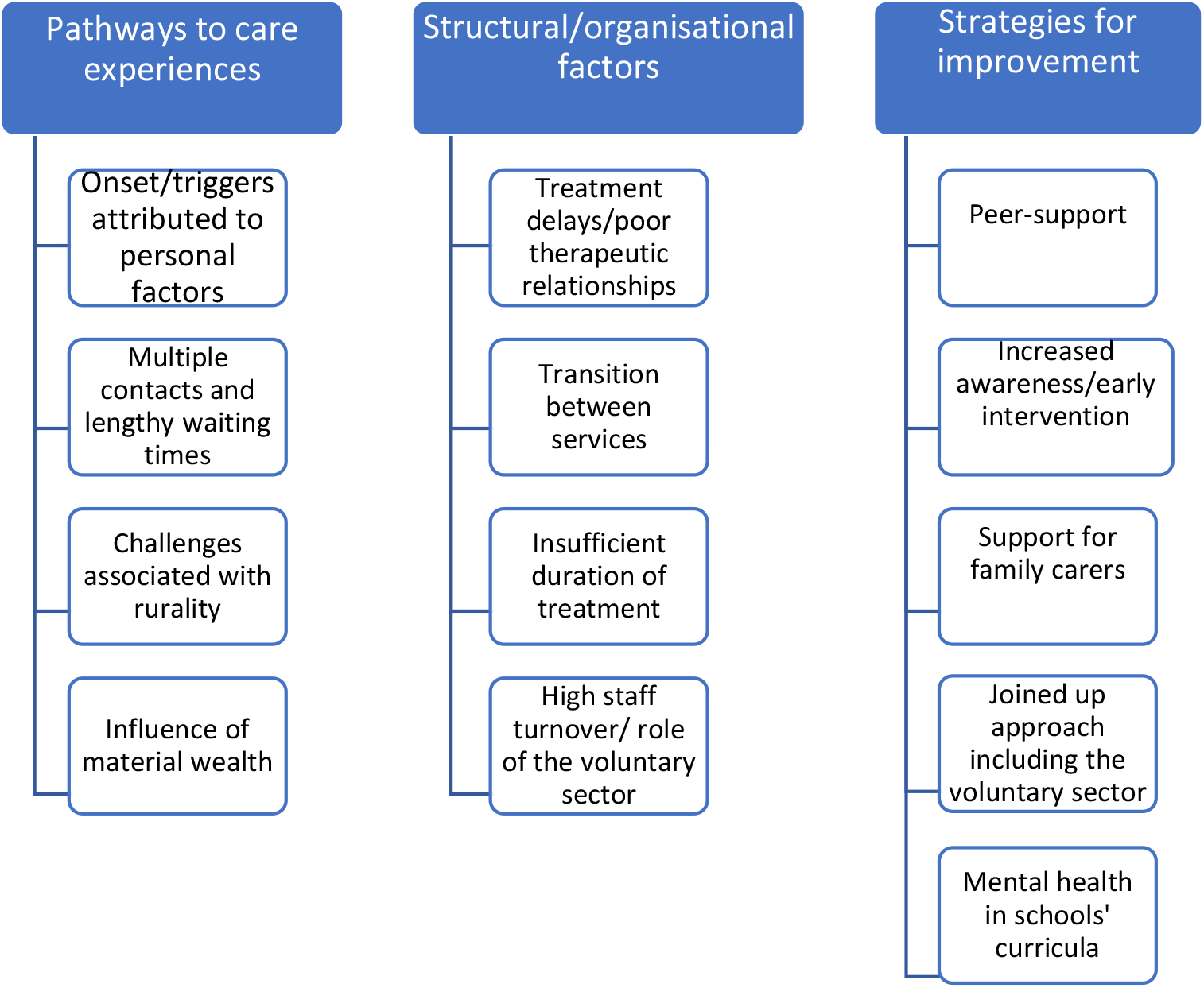
A summary of the overarching themes and sub-themes.

### Pathways to care experiences

#### Onset or triggers of mental health difficulties

Young people and family carers all reported noticing changes in their own or their child’s mental health during secondary school, and many described a gradual onset of symptoms. They attributed onset of difficulties to personal factors such as shyness, sexuality, dyslexia, or life events such as bullying and exams. All FCs reported lacking mental health knowledge and feeling under-equipped to deal with their child’s mental health difficulties.

> *YP 3: But I think just a lot of past things that happened, I kind of triggered, kind of just becoming anxious and not really seeing like, the way forward and things*.

> *FC 1: But it was a lack of knowledge on my part of understanding about that, and the impact for him. And, you know, since I’ve obviously tried to learn about stuff, educate myself understand things from his point of view, but I just didn’t have a clue*.

#### Multiple contacts and lengthy waiting times

In terms of help-seeking, all YP and FC participants reported the benefit of having more than one key contact in their pathways to care. Help was mostly sought from general practitioners (GPs), although most participants described challenges in accessing timely support due to delays in GP referrals and the long waiting times for mental health services. Three participants (2FCs, 1YP) reported they sought help directly from mental health services during crises (e.g., self-harm & suicidal ideation).

> *VOS: I could not be more disappointed in the services; I could not be more unhappy with what I’ve seen*.

> *YP 4: I mean for starters it took like eight months for me to actually get someone and realistically, when you need help you need it right then, and waiting for a long period of time makes it so much worse*.

> *FC 2: We were using the crisis team. And she was basically on 24/7 watch. And they were seeing her or talking to her most days. Very, very scary time*.

#### Material wealth and socioeconomic status

Linked to the lengthy waiting times for accessing mental health services, five participants (2FCs, 3YP) sought help from private healthcare providers. However, they recognised that not everyone could afford private treatment and highlighted the urgency in increasing mental health service capacity.

> *FC 4: And it was apparent, there was a massive waiting list. And we didn’t try too hard. We just thought actually, let’s not bother. We’re losing time. We’ll access that (private clinic). It’s through the company, my husband works at*.

> *YP 1: But I went into the private sector afterwards. And the person I had was brilliant*.

#### Challenges associated with rurality

Most of the participants did not feel living in a rural area impacted their own ability to access mental health support, given they had the resources and means to travel to mental health clinics. However, all but one acknowledged the potential challenges that rurality could pose for accessing support without socioeconomic means (e.g., car ownership), which in turn could influence treatment delays and disengagement with services. Stigma was also identified as a barrier to help seeking

> *FC 2: Obviously, if I didn’t, you know, if we didn’t drive, then obviously, it would have been a problem getting her to appointments and stuff*.

> *YP 3: For me to access care. It was a 30-minute drive. And obviously, yeah, if you don’t have parents who drive you can’t access it. The bus goes to the bus station, and you’d have to get another one then you won’t get there on time for your appointment. So, I think it is just being aware of how far away they are*.

> *FC 1: because of the stigma around mental health, there wasn’t many people like outside of my immediate family in the household, I didn’t talk to anyone about what was going on with my son. So, to respect his privacy, it becomes a bit of a secret*.

#### Facilitators

Despite the barriers discussed above, several participants identified key enablers that helped them in their help-seeking and access to mental healthcare. These included family involvement/support, school nurses, being listened to, and shorter delays in accessing specialist services such as eating disorder or early-intervention psychosis services.

> *YP 4: Yeah, I think my mom is quite good at, like, telling when something’s wrong. Even if we don’t even recognise it.*

> *YP 3: Someone just simply saying, you know, if you need to talk about it, I’m here type of thing*

### Structural/organisational factors

Practical barriers that delayed or hindered treatment emerged in five participants (3FC, 2YP). These included limited therapy session numbers, referral between services, problematic transitions from children to adult services and poor therapeutic relationships.

#### Poor therapeutic relationships

Two FC participants recounted negative experiences of telephone-based assessments, which led their child to disengage with the services.

> *FC 3: I think the very first psychiatry appointment was over the phone. And that was just awful, because she’d never met (daughter)*.

> *FC 1: And I was so excited about NHS 111. Number two option, I thought, oh, yeah, it sounds fantastic. They didn’t seem to know what to do*.

Two FC participants described high staff turnover as detrimental to their child’s recovery.

> *FC 2: The only thing I would say is that they are so overworked and under supported, that they don’t last very long. So, it’s hindered (daughter’s) progress, because it keeps changing. The family therapist’s left. And we weren’t offered an alternative*.

> *FC 3: She fell out with one of her workers. And then another one left. So, she’s seen an awful lot of people. And obviously, she has to keep repeating stuff that she knows is very hard for her to talk about*.

#### Transition between services

Three FC participants described a sense of hopelessness in statutory services especially when their children turned 18 years old and needed to transition to adult services. They felt the transition was disjointed and one FC reported her child was discharged to the GP, as there was lack of capacity in adult mental health services.

> *FC 3: And then of course, the sad thing is that you know, you turn 18. And yeah, everything just stops overnight*.

> *FC 4: And then with a psychiatrist, it was literally just one appointment. And she said, “Well, I’m handing it over to a GP now, because you’re 18”*.

#### Insufficient duration of treatment

Many participants described negative experiences related to the treatment they received from mental health services. Evidence emerged in four transcripts that the number of therapy sessions offered were insufficient, especially as many reported there were numerous past events that needed ‘unpicking’. This is also linked to the decision some made to pay for private therapy/treatment.

> *YP 4: …and they only give you around six weeks of someone to talk to and it’s only for like 40 minutes. That’s just not enough*.

> *VOS: It’s CBT for five to eight sessions or nothing. And it’s on the ground, like on a superficial level. It’s not enough. There’s not enough there*.

#### The role of voluntary organisations

There was evidence of limited awareness of mental health support available from voluntary organisations. One FC participant recalled a negative experience when her daughter was referred by the statutory services to a charity organisation for additional support. Another FC participant recognised the positive difference the third sector could make in supporting young people with mental health difficulties. However, she highlighted that lack of investment impedes the sector’s service coverage.

> *FC 1: And I know that we will be redirected to the third sector organisation who are lovely but have a waiting list*.

#### Facilitators

Some participants reported positive experiences in the treatment and care they received. One reported their child received timely support during a crisis, although she attributed this to her child experiencing an eating disorder. Another FC participant described a positive experience in receiving family therapy alongside her child’s treatment.

> *FC 3: We’ve had family therapies; we were offered family therapy. So, the care we’ve received, it’s been fantastic*.

> *FC 2: And I’ve been told, because it’s an eating disorder that they have to be seen, so they get sort of pushed to the top of the list*.

### Strategies for improvement

Participants offered several suggestions about how to improve pathways to care and future help-seeking experiences for young people in general and for those living in rural areas. These include: (a) support for family e.g., emotional support; (b) increasing public awareness about young people’s mental health; (c) more investment in mental health provision; (d) embedding mental health in schools’ curricula; (e) early intervention and more joined up working, including the third sector; (f) employing and training young people to support other young people with a clear pathway for them to access professional support if needed; (g) community champions in schools and villages; (h) in-person contact rather than virtual. Table 2 provides illustrative quotations from participants.

**Table 2:**
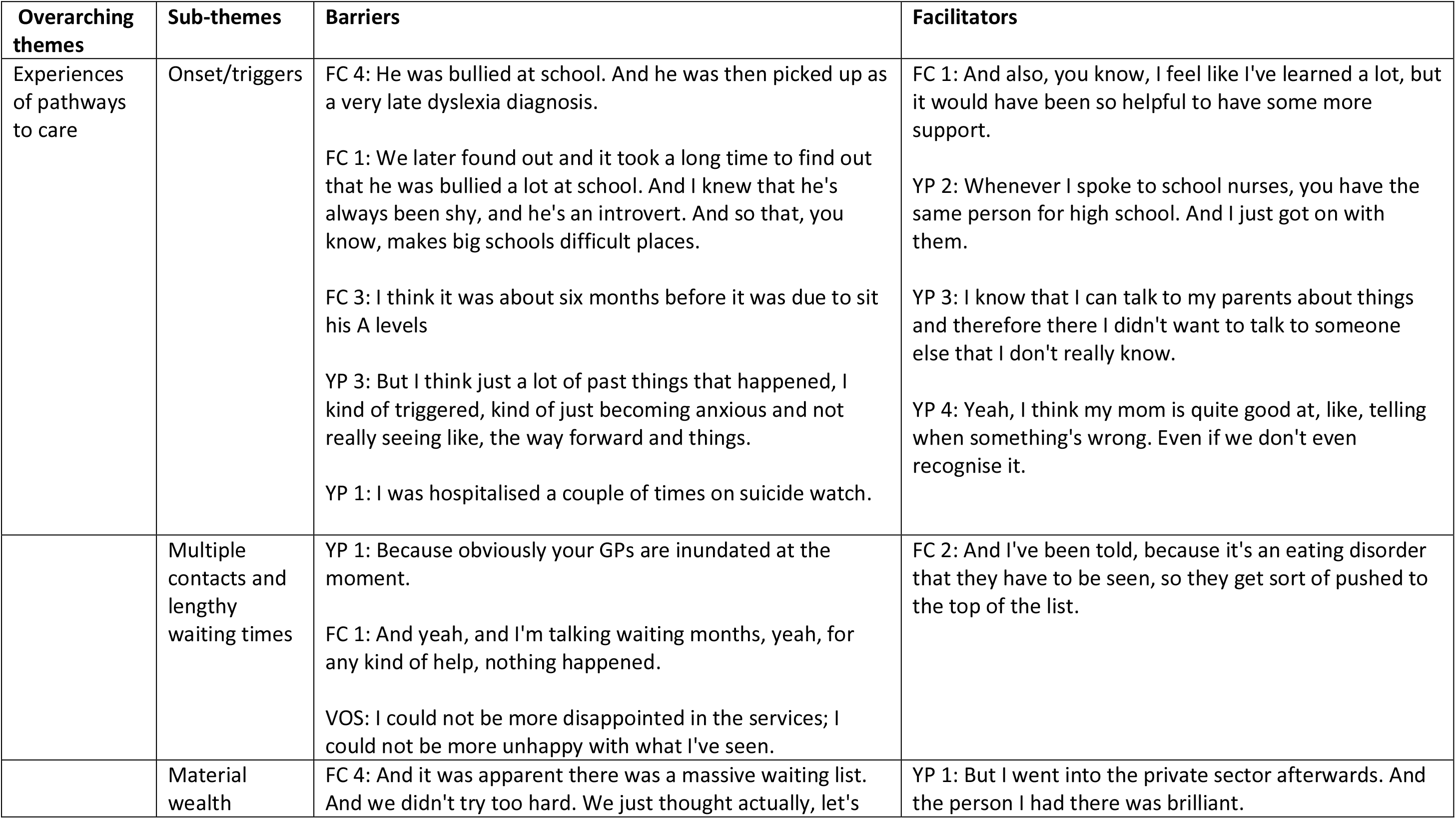
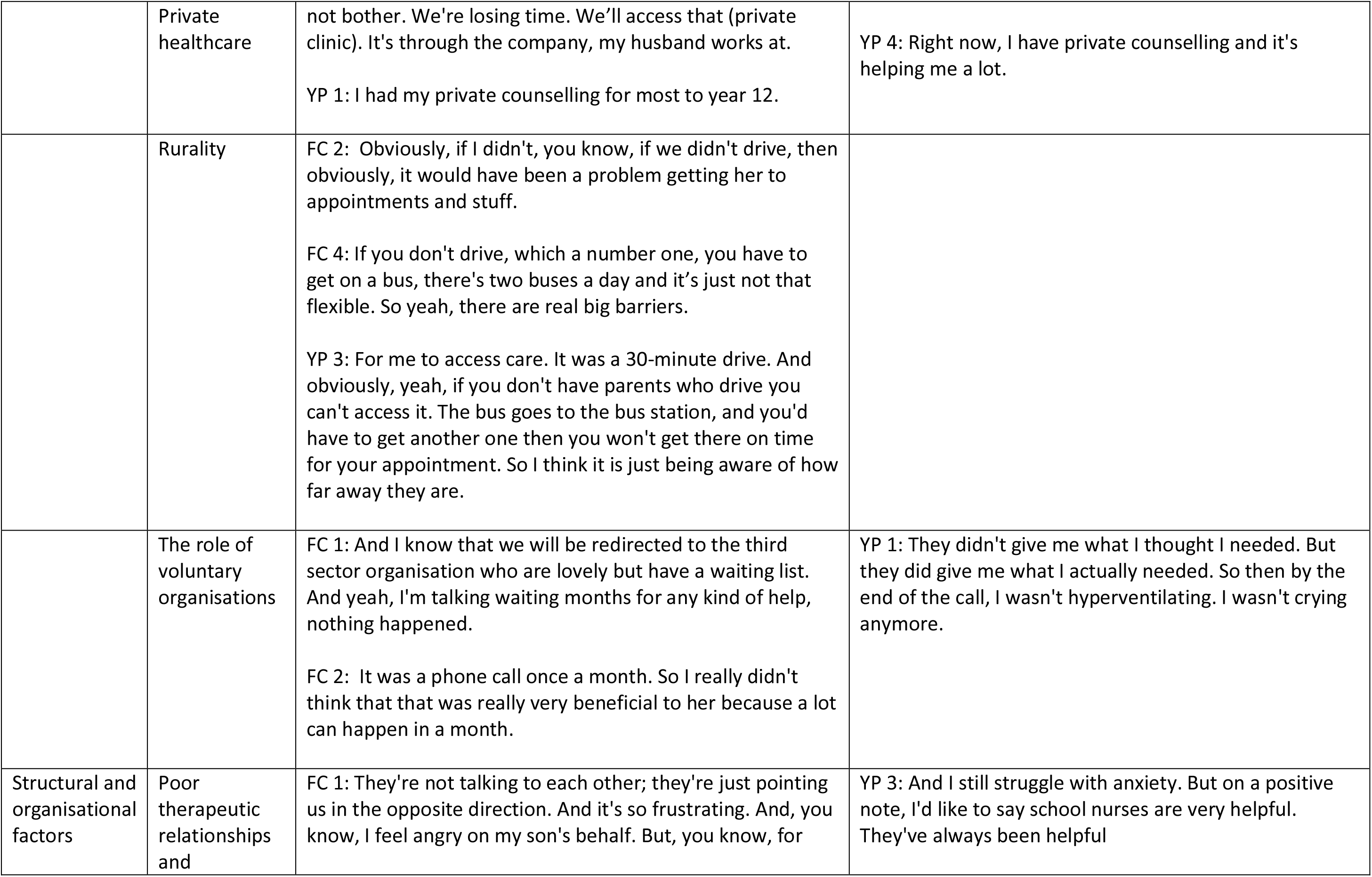
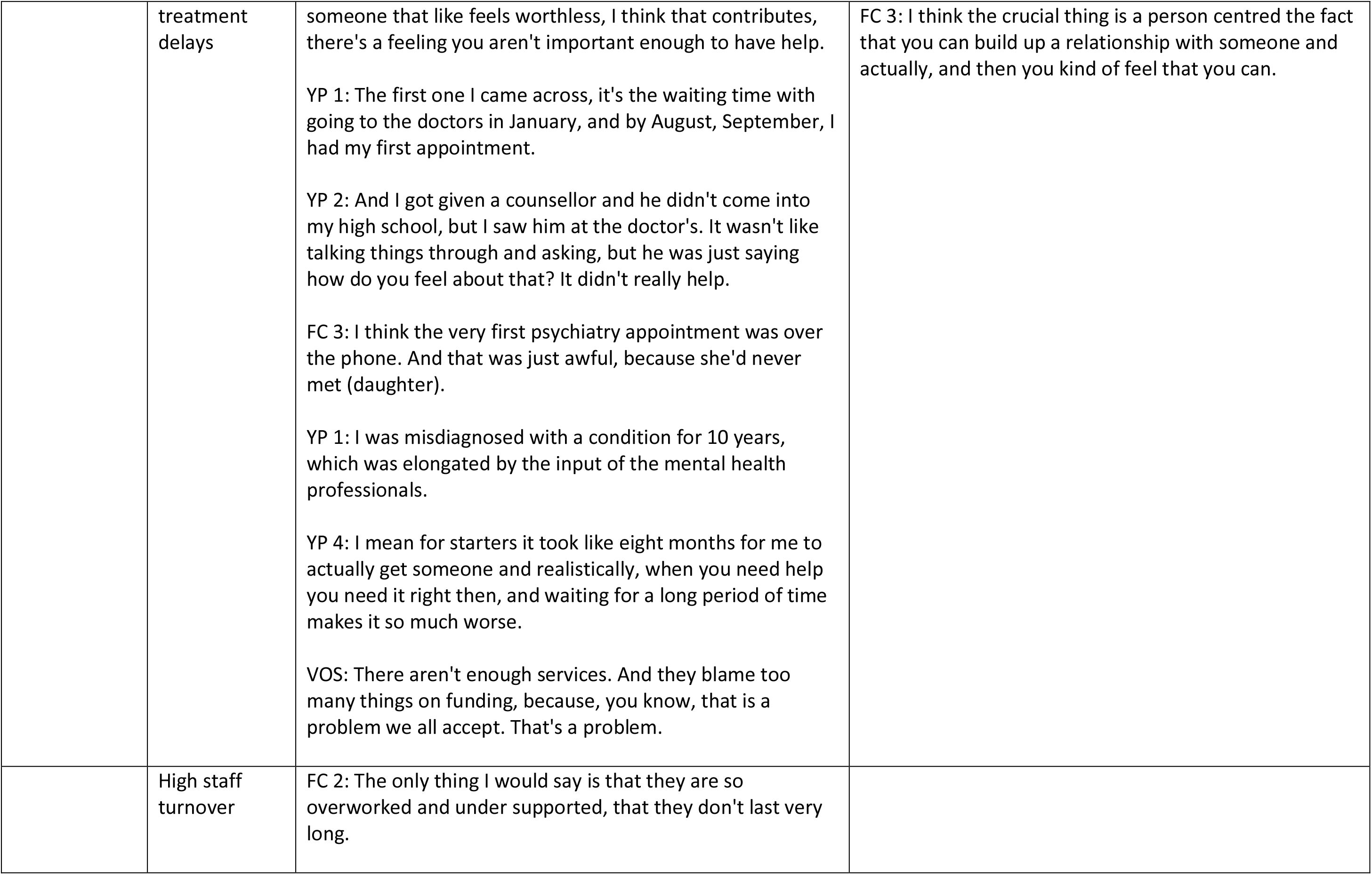
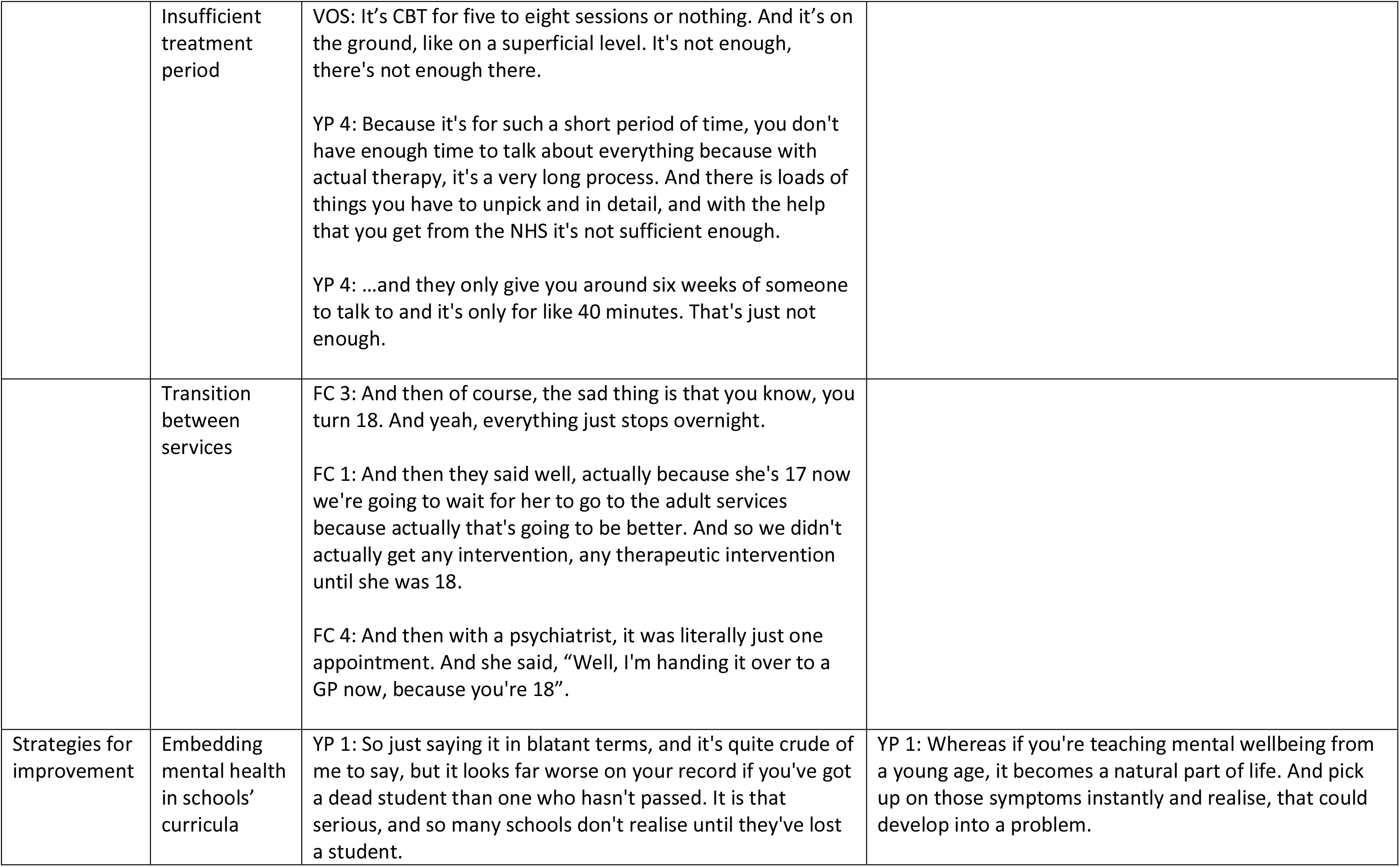
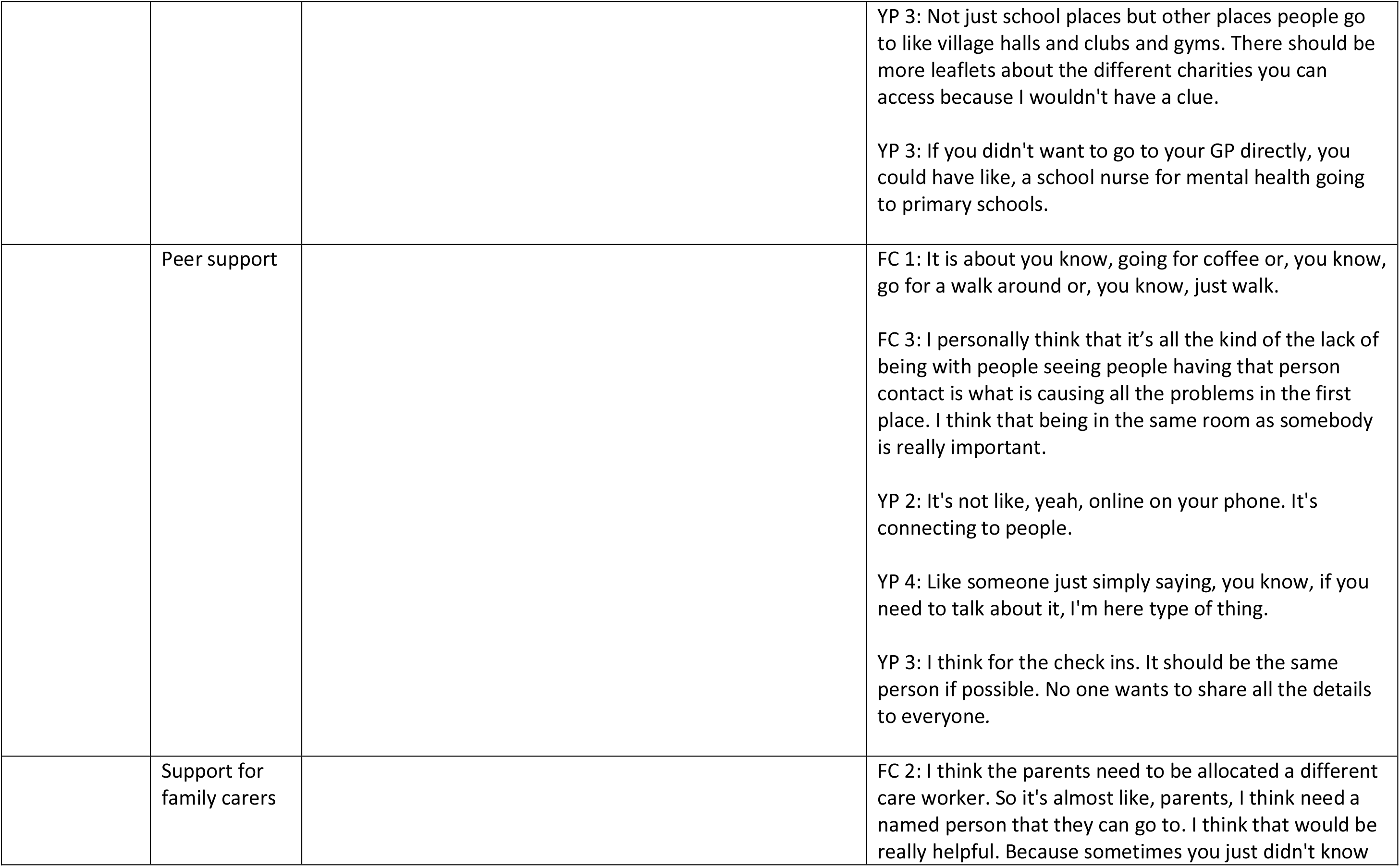
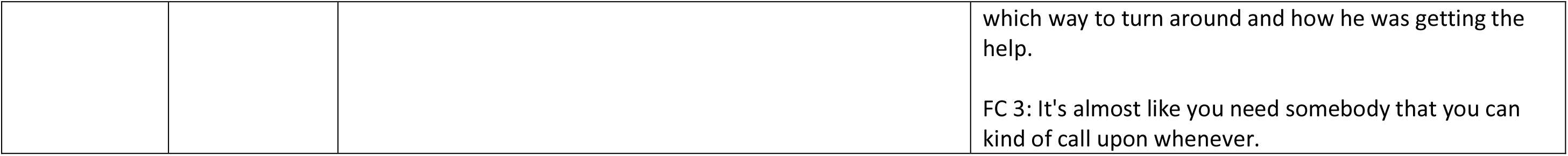
Illustrative quotations of barriers and facilitators of help-seeking for mental health difficulties.

## DISCUSSION

### Summary of findings

Our data suggest there are several barriers related to help-seeking for mental health difficulties among young people living in rural areas, including lengthy waiting times, material wealth, stigma, poor therapeutic relationships with healthcare providers, insufficient treatment duration, poor transition between services, high staff turnover and limited investment in mental health services. Our participants highlighted that people with lower socioeconomic status (e.g., a car or money for private treatment) are likely to be most affected by rurality. Findings from this exploratory work suggest there are several avenues to pursue in future research aimed at improving access to mental healthcare in rural areas, including early intervention, peer-support, support for carers, and increasing mental health awareness in schools and communities.

### Relationship of findings to previous research

Only a handful of studies have been undertaken in the UK exploring help-seeking for mental health difficulties in UK rural populations. The significance of family and friends in help-seeking for mental health difficulty was shown in a UK study by Allan and colleagues (*under review*) who interviewed 11 young people with at-risk-mental-state and first episode psychosis in East of England, UK, about barriers and facilitators of help-seeking. They found that nine of the eleven participants reported friends and family as key facilitators of timely access to treatment (Allan, under review). Of the available international studies, the potential impact of socioeconomic status in help-seeking among rural populations was shown in an Australian study by Boyd et al. (2011) who found that difficulties associated with travelling to obtain help were significant concerns for young people in rural areas (Boyd et al., 2011).

Our findings of the importance of lengthy waiting times, material wealth and organisational factors are consistent with a Canadian study by Boydell et al. (2006), who conducted semi-structured interviews with 30 family members and 35 young people, and reported three overarching themes (personal, systemic and environmental) that influenced their participants’ ability to navigate the mental healthcare system (Boydell et al., 2006). Our findings of help-seeking during a crisis are echoed in USA and Australian studies, where Pisani and colleagues (2012) and Boyd et al. (2011) reported that young people in their studies mostly sought professional help seeking for suicidal ideations and severe anxiety (Boyd et al., 2011; Pisani et al., 2012). However, in contrast to our results, Thompson et al. (2018) investigated help-seeking behaviour among adolescents and young adults in a USA study using Crisis Text Line, they found rurality as a strong predictor of low rate of help-seeking, despite elevated rates of suicide among their rural participants (Thompson et al., 2018).

Our results regarding poor therapeutic relationships and insufficient treatment duration are consistent with several studies. For example, Russell and colleagues (2004) explored help-seeking barriers faced by young men with suicidal ideations in a rural Irish setting; they interviewed 71 young men and 79 key informants, and their participants reported that professional help was unacceptable and that friends and family were their preferred sources of support (Russell et al., 2004). Another study from Australia in which Boyd et al. (2007) investigated the perspectives of young people in help-seeking for mental health problems in rural areas showed that lack of information about rural services and seeking help from a GP impeded their access to appropriate care (Boyd et al., 2007). Furthermore, our findings of multiple contacts in help-seeking and stigma were reflected in several other studies (Boyd et al., 2007; Bradley et al., 2010; Sears, 2004).

In the UK, mental healthcare is free on the National Health Service (NHS). According to the NHS Long-term Plan, the UK government pledges a commitment to ensure that people’s health is not compromised because of (a) where they live, (b) what services and treatment they can access, and (c) because they do not have access to financial resources (NHS England, 2019). However, our findings highlight serious inequalities for young people in accessing mental health support, and so more work and investments are needed to achieve the Long-term Plan pledges.

### Methodological considerations: strengths and limitations

To our knowledge, this is the first study to interview different stakeholder groups to understand challenges and opportunities in care pathways for mental health difficulties in rural populations in the UK. Our qualitative approach meant we were able to explore in detail the existence and mechanisms underlying varied barriers and facilities of care pathways amongst our sample. This allowed us to report rich findings and recommendations for future work. In particular, the applied nature of our topic guides means that there are important implications for NHS service commissioning and provision. An additional strength is the varied perspectives of family carers, young people with lived experience, and representation of the voluntary sector, given many previous international studies have largely reported findings solely from patient or young people’s perspectives.

There are some limitations to consider when interpreting the study findings. First, the study was limited by the small sample size, particularly, as only one voluntary sector worker participated. Second, the challenges to recruit people from the most remote areas meant their views may not be represented. Their inclusion could have provided a more rounded picture of the scale and extent of challenges of mental health care provision in rural and remote populations. Third, our sample was quite homogenous in terms of age, gender, ethnicity, and socioeconomic status, and therefore the participants’ views may not reflect the experiences of minority groups living in rural areas. Future research would benefit from exploring barriers and facilitators to help-seeking for other demographic groups such as people from minority ethnic groups, LGBTQ+, young men, and individuals living in the most deprived rural areas.

### Clinical implications and interpretation of findings

There are several key findings from our study that would be beneficial to clinical practice and future research. Our findings show that:

- Pathways into mental health care are complex and involve multiple individuals and organisations, including family members, GPs, schools, and voluntary organisations. Therefore, interventions to improve access to mental health care should be systemic in nature and consider the varied and interacting roles of all stakeholders.
- The impact of rurality on access to mental health care is more nuanced than geographical location and distance from services. Stigma and access to socioeconomic means to overcome rural barriers (e.g., a car, money for private treatment) are important factors of rurality in seeking help for mental health difficulties. This intersection requires further investigation.
- Family involvement and support was identified as an important facilitator of successful help seeking. Whilst mental health care often focuses on the individual, providing education and supportive interventions for family members may aid early identification of mental health difficulties.
- Whilst online interventions may be a potential strategy to improve the reach of mental healthcare, particularly in rural communities, many participants expressed a preference for in-person contact.
- There are potential strengths in rural communities due to their active community presence. Therefore future interventions might include training young people to support others within their own community, and identifying community mental health champions.
- More investment in mental health service provision is required. Whilst community-level interventions can improve pathways into care, they are not a replacement for specialist mental health services.

## CONCLUSIONS

This qualitative study showed that young people, relatives, and voluntary organisation workers find accessing care and support for mental health problems challenging, and they report the need for an intervention to reduce the distress. Training young people to support other young people; and identifying mental health champions in rural communities could reduce distress and ameliorate pressures on the health service.

## Data Availability

All data produced in the present work are contained in the manuscript

## Acknowledgements

We thank all those who took part in the study and shared their experiences.

## Authors’ contributions

SO conceived the idea of the study. All authors were involved in the design of the study, and all were involved in the data collection and data analysis. SO prepared the manuscript with input from all authors. All authors were involved in the interpretation of the data and in commenting on and revising drafts of the paper.

## Ethical approval

Ethical approval for they study was obtained from the University of East Anglia, Faculty of Medicine & Health Sciences Research Ethics Committee (Reference: 2021/22-018).

## Informed Consent

All participants were provided with information about the study, offered an opportunity to speak with the researchers prior to taking part, and gave written consent for both participation in interviews and use of anonymised quotes for publication.

## References

Allan, S., Oduola, S., Ricco, S., & Jodgekins, J. (under review). “Jumping from place to place”: Service User Perspectives on Pathways to care in At-Risk Mental States and First Episode Psychosis. Psychosis.

Allan, S. M., Hodgekins, J., Beazley, P., & Oduola, S. (2020). Pathways to care in at-risk mental states: A systematic review. Early Interv Psychiatry. 10.1111/eip.13053

Allwood, L. (2021). The space between us: Children’s mental health and wellbeing in isolated areas.

Centre for Mental Health. https://www.centreformentalhealth.org.uk/sites/default/files/publication/download/CentreforMH_TheSpaceBetweenUs_Rurality.pdf

Andrilla, C. H. A., Patterson, D. G., Garberson, L. A., Coulthard, C., & Larson, E. H. (2018). Geographic Variation in the Supply of Selected Behavioral Health Providers. Am J Prev Med, *54*(6 Suppl 3), S199-s207. 10.1016/j.amepre.2018.01.004

Bhui, K., Ullrich, S., & Coid, J. W. (2014). Which pathways to psychiatric care lead to earlier treatment and a shorter duration of first-episode psychosis? BMC Psychiatry, 14, 72. 10.1186/1471-244x-14-72

Boyd, C., Francis, K., Aisbett, D., Newnham, K., Sewell, J., Dawes, G., & Nurse, S. (2007). Australian rural adolescents’ experiences of accessing psychological help for a mental health problem. Aust J Rural Health, 15(3), 196–200. 10.1111/j.1440-1584.2007.00884.x

Boyd, C. P., Hayes, L., Nurse, S., Aisbett, D. L., Francis, K., Newnham, K., & Sewell, J. (2011). Preferences and intention of rural adolescents toward seeking help for mental health problems. Rural Remote Health, 11(1), 1582.

Boydell, K. M., Pong, R., Volpe, T., Tilleczek, K., Wilson, E., & Lemieux, S. (2006). Family perspectives on pathways to mental health care for children and youth in rural communities. J Rural Health, 22(2), 182–188. 10.1111/j.1748-0361.2006.00029.x

Bradley, K. L., McGrath, P. J., Brannen, C. L., & Bagnell, A. L. (2010). Adolescents’ attitudes and opinions about depression treatment. Community Ment Health J, 46(3), 242–251. 10.1007/s10597-009-9224-5

Braun, V., & Clarke, V. (2006). Using thematic analysis in psychology. Qualitative Research in Psychology, 3(2), 77–101. 10.1191/1478088706qp063oa

Costas, M. (2017). Improving mental health services. Rural Mental Health Matters. Retrieved September 2021 from https://www.ruralmentalhealthmatters.co.uk/

Dept. for Environment Food & Rural Affairs. (2021). Statistical Digest of Rural England: Population Retrieved June 2022 from https://assets.publishing.service.gov.uk/government/uploads/system/uploads/attachment_data/file/1028819/Rural_population__Oct_2021.pdf

Dhakal, K. (2022). NVivo. Journal of the Medical Library Association: JMLA, 110(2), 270.

Edrawsoft. (2022). Mind Map Software https://www.edrawsoft.com/free-mind-map-tool.html

Ford, T., John, A., & Gunnell, D. (2021). Mental health of children and young people during pandemic. Bmj, 372, n614. 10.1136/bmj.n614

Hodgekins, J., Clarke, T., Cole, H., Markides, C., Ugochukwu, U., Cairns, P., Lower, R., Fowler, D., & Wilson, J. (2017). Pathways to care of young people accessing a pilot specialist youth mental health service in Norfolk, United Kingdom. Early Interv Psychiatry, 11(5), 436–443. 10.1111/eip.12338

Kirkbride, J. B., Hameed, Y., Ankireddypalli, G., Ioannidis, K., Crane, C. M., Nasir, M., Kabacs, N., Metastasio, A., Jenkins, O., Espandian, A., Spyridi, S., Ralevic, D., Siddabattuni, S., Walden, B., Adeoye, A., Perez, J., & Jones, P. B. (2016). The Epidemiology of First-Episode Psychosis in Early Intervention in Psychosis Services: Findings From the Social Epidemiology of Psychoses in East Anglia [SEPEA] Study. Am J Psychiatry, appiajp201616010103. 10.1176/appi.ajp.2016.16010103

Lewinsohn, P. M., Rohde, P., & Seeley, J. R. (1998). Major depressive disorder in older adolescents: prevalence, risk factors, and clinical implications. Clin Psychol Rev, 18(7), 765–794. 10.1016/s0272-7358(98)00010-5

MacDonald, K., Ferrari, M., Fainman-Adelman, N., & Iyer, S. N. (2021). Experiences of pathways to mental health services for young people and their carers: a qualitative meta-synthesis review. Soc Psychiatry Psychiatr Epidemiol, 56(3), 339–361. 10.1007/s00127-020-01976-9

Morales, D. A., Barksdale, C. L., & Beckel-Mitchener, A. C. (2020). A call to action to address rural mental health disparities. J Clin Transl Sci, 4(5), 463–467. 10.1017/cts.2020.42

NHS England. (2019). The NHS Long Term Plan. https://www.longtermplan.nhs.uk/areas-of-work/mental-health/

NHS Providers. (2019). NHS Mental health implementation plan. Retrieved 16/04/2020 from https://www.longtermplan.nhs.uk/wp-content/uploads/2019/07/nhs-mental-health-implementation-plan-2019-20-2023-24.pdf

Onwumere, J., Zhou, Z., & Kuipers, E. (2018). Informal Caregiving Relationships in Psychosis: Reviewing the Impact of Patient Violence on Caregivers. Front Psychol, 9, 1530. 10.3389/fpsyg.2018.01530

Pisani, A. R., Schmeelk-Cone, K., Gunzler, D., Petrova, M., Goldston, D. B., Tu, X., & Wyman, P. A. (2012). Associations between suicidal high school students’ help-seeking and their attitudes and perceptions of social environment. J Youth Adolesc, 41(10), 1312–1324. 10.1007/s10964-012-9766-7

QSR International Pty Ltd. (2020). NVivo 20. https://www.qsrinternational.com/nvivo-qualitative-data-analysis-software/home

Russell, V., Gaffney, P., Collins, K., Bergin, A., & Bedford, D. (2004). Problems experienced by young men and attitudes to help-seeking in a rural Irish community. Ir J Psychol Med, 21(1), 6–10. 10.1017/s0790966700008065

Sears, H. A. (2004). Adolescents in rural communities seeking help: who reports problems and who sees professionals? J Child Psychol Psychiatry, 45(2), 396–404. 10.1111/j.1469-7610.2004.00230.x

The King’s Fund. (2018). Funding and staffing of NHS mental health providers: still waiting for parity. Retrieved June 2022 from https://www.kingsfund.org.uk/publications/funding-staffing-mental-health-providers

Thompson, L. K., Sugg, M. M., & Runkle, J. R. (2018). Adolescents in crisis: A geographic exploration of help-seeking behavior using data from Crisis Text Line. Soc Sci Med, 215, 69–79. 10.1016/j.socscimed.2018.08.025

WHO. (2018). The Global Health Observatory. Retrieved December 2021 from https://www.who.int/data/gho/data/themes/mental-health

